# Health Economic Evaluation of a Double-Blind, Randomised, Placebo-Controlled Trial of Low-Dose Oral Morphine (MABEL)

**DOI:** 10.1101/2025.06.04.25328980

**Authors:** M Atter, PS Hall, RA Evans, J Norrie, J Cohen, B Williams, N Chaudhuri, S Bajwah, IJ Higginson, M Pearson, DC Currow, G Stewart, MT Fallon, MJ Johnson

**Affiliations:** Edinburgh Clinical Trials Unit, The Usher Institute, University of Edinburgh; Department of Respiratory Sciences, NIHR Leicester Biomedical Research Centre – Respiratory, University of Leicester; Hull Health Trials Unit, Hull York Medical School, University of Hull; School of Medicine, Department of Life and Health Sciences, Ulster University; Cicely Saunders Institute, King’s College London; Faculty of Health, University of Technology Sydney, Sydney, NSW, Australia; Western General Hospital, NHS Lothian

**Keywords:** Economics, Clinical Trial, Morphine, Respiration

## Abstract

**Objectives:** To compare costs and health consequences and to assess the cost-effectiveness of using low-dose oral long-acting morphine in people with chronic breathlessness.

**Design:** Within-trial planned cost-consequences and cost-effectiveness analysis of data from a multi-site, parallel-group, double-blind, randomised, placebo-controlled trial of low-dose, long-acting morphine.

**Setting:** 11 hospital outpatients across the UK.

**Participants:** Those eligible to participate were consenting adults with chronic breathlessness due to long-term cardiorespiratory conditions.

**Intervention:** 5-10mg twice-daily oral long-acting morphine with a blinded laxative for 56 days.

**Primary outcome measures:** Mean and standard deviation (SD) of healthcare resource use (HRU) by trial arm; mean differences and 95% confidence intervals (CI) of costs between trial arms.

**Secondary outcome measures:** Mean differences in 28- and 56-day quality-adjusted life years (QALYs based on EQ-5D), SF-6D scores, and ICECAP-SCM scores; cost-utility of long-acting morphine for chronic breathlessness.

**Results:** 143 participants (75 morphine, 67 placebo) were randomised; 140 formed the modified intention-to-treat population (90% power; males 66%; mean age 70.5 [SD 9.4]). There were more inpatient and fewer outpatient services used by the morphine group *versus* placebo. In the base case analysis at 56 days, long-acting morphine was associated with similar mean per-patient costs and QALYs: There was an increase of £24 (95% CI: -£395, £552) and 0.002 (95% CI: -0.004, 0.008) QALYs. Hospitalisations were the main driver of cost differences. The corresponding incremental cost-effectiveness ratio (ICER) was £12,000/QALY, with a probability of cost-effectiveness of 54% at a £20,000 Willingness-to-Pay (WTP) threshold. In the scenario analysis that excluded costs of adverse events considered unrelated to long-acting morphine by site investigators and researchers, the probability of cost-effectiveness increased to 73%.

**Conclusion:** Oral morphine for chronic breathlessness is likely to be a cost-effective intervention provided adverse events are minimised but the effect on outcome is small and cautious interpretation is warranted.

**ARTICLE SUMMARY:** *Strengths and limitations of this study:*

- Comprehensive collection of patient-reported health economic data in a randomised controlled trial, including three different health outcome measures relevant to people living with chronic breathlessness due to medical conditions
- The parent trial intervention dosing schedule mirrored clinical practice to give a pragmatic indication of cost impacts
- Limited interpretation of cost-effectiveness analysis due to a likely random imbalance in deaths and expensive adverse events, in a study design not primarily designed to detect differences in economic endpoints
- Technical challenges in analysing and estimating costs for concomitant medications

## INTRODUCTION

Chronic breathlessness is a disabling symptom common in cardio-respiratory diseases and cancer. [1,2] Breathlessness is a global problem, with population prevalence ranging from 9% in high-income countries to 40% in low-income countries such as India, mirroring countries with higher rates of causal diseases, higher smoking rates and greater levels of environmental pollution. [3,4] Despite the significant adverse impacts of chronic, or persistent, breathlessness on quality of life, physical and psychosocial functioning, and healthcare service utilisation, effective management is often neglected. [5–7]

Opioids have shown potential in reducing the perception of breathlessness by modulating central pathways, but clinical trials in ambulant populations have not consistently demonstrated benefits, partly due to short study durations and the lack of measures of exercise endurance. In addition, morphine has a well-known adverse effect profile, including complications which could plausibly increase health service costs, complications from gastrointestinal and neurocognitive side effects, and respiratory depression. The health economic implications of chronic breathlessness beyond the specific context of chronic obstructive pulmonary disease (COPD) have not been comprehensively studied. The only published health economic evaluation, to our knowledge, however, found a high likelihood that four weeks of long-acting morphine was cost-effective, compared with placebo, in the management of people living with chronic breathlessness due to COPD. [7]

The Morphine And BrEathLessness (MABEL) trial aimed to assess the effectiveness, cost-consequences, cost-effectiveness and safety of long-acting low-dose, oral long-acting morphine on patient-reported worst breathlessness in people with chronic breathlessness. [8]

This paper contains the health economic analysis component of the MABEL trial. This analysis aimed to compare the costs, health outcomes (cost-consequences analysis), and cost-effectiveness of using morphine on patient-reported *worst breathlessness* in people with chronic breathlessness.

## METHODS

### Trial overview

Full details of the MABEL trial, its procedures (including inclusion/exclusion criteria), and clinical findings can be found in the study protocol and main results paper; this section’s summary is included for context. [8]

MABEL was an 11-site, UK-based multicentre, Phase-III, parallel-group, double-blind, randomised placebo-controlled titration trial. Eligible patients were ambulant adults with moderate to severe chronic breathlessness (mMRC ≥ grade 3 or 4) due to cardio-respiratory disease or cancer, with adequate renal function, who gave informed consent. The *Active* (Morphine) intervention consisted of 5-10mg twice-daily oral long-acting morphine with a blinded laxative, for 56 days. The primary outcome was worst breathlessness in the past 24 hours (measured by a 0-10 numerical rating scale (NRS) on Day 28. Outcome measures pertinent to the health economic evaluation were the Short-form-12 (SF-12), the EuroQol-5dimension-5level score (EQ-5D-5L), ICEpop CAPability-Supportive Care Measure (ICECAP-SCM) and Health Resource Use Questionnaire measured at baseline, Day 28 and Day 56. [9–12] Attribution of serious adverse events to study drug was assigned by the recruiting site clinician, and discussed with the clinical Chief-Investigator (with over thirty years’ experience of opioid prescribing).

### Methods for the economic analysis

The paper follows Consolidated Health Economic Evaluation Reporting Standards (CHEERS) guidance for health economic evaluations (summarised in Supplementary Table 1). [13] The methods of calculating costs, health outcomes and cost-effectiveness metrics were stipulated in a Health Economic Analysis Plan (HEAP) approved by the lead economist and chief investigator before the data lock and unblinding (see Supplementary Materials).

In line with guidance, all costs and health outcomes were calculated from a National Health Service (NHS) & Personal Social Services perspective, which includes the costs of primary and secondary care, and community NHS activity in England. [14]

As per the National Institute for Health and Care Excellence (NICE) guidelines, the health outcome of interest is the quality-adjusted life years (QALY), which is an “index of survival that is adjusted to account for the patient’s quality of life” best calculated using the EQ-5D-5L measure. [14,15]

In light of the evolving discussion in the scientific literature about the interpretation of the QALY in end-of-life settings, SF-6D (derived from SF-12) and the capability measure the ICECAP-SCM were selected as alternative measures in the sensitivity analyses. [9,10]

### Data collection

The data needed for the health economic analysis were drawn from a combination of trial case report form (CRF) data and a modified version of the UK Cancer Costs Questionnaire collected on Day 28 and Day 56 after randomisation. [12] The time horizon used in this paper is therefore 56 days, but selected key results for Day 28 are also presented. With a time horizon of under 1 year, no discounting of costs or outcomes were applied.

### Estimating outcomes

QALYs were calculated individually for each patient in the intention-to-treat population by using the following steps: First calculating Health State Utility Values (HSUVs) from EQ-5D-5L Patient-Reported Outcome Measure (PROM) questionnaire data at each time point using a validated mapping function by Hernández Alava et al. [16] subsequently calculating 56-day QALYs as a function of HSUVs and their corresponding time points using the area-under-the-curve method outlined by Manca et al. [17]

The SF-6D was converted into HSUVs using validated ProCore software provided by QualityMetric, while the ICECAP-SCM measure was converted into tariff values for its capability score, which ranges from 0 (no capability) to 1 (full capability), using its published value set. [18,19] SF-6D and ICECAP-SCM scores were not used in QALY calculation or the cost-effectiveness analysis.

### Estimating costs

Price weights used to calculate per-patient costs of hospital and community (hospice) resource use are presented in Table 1. Medication unit costs, which include morphine as the investigational medicinal product (IMP), laxatives as the non-investigational medicinal product (NIMP), and concomitant medications, were sourced from the British National Formulary (BNF). [20]

**Table 1:**
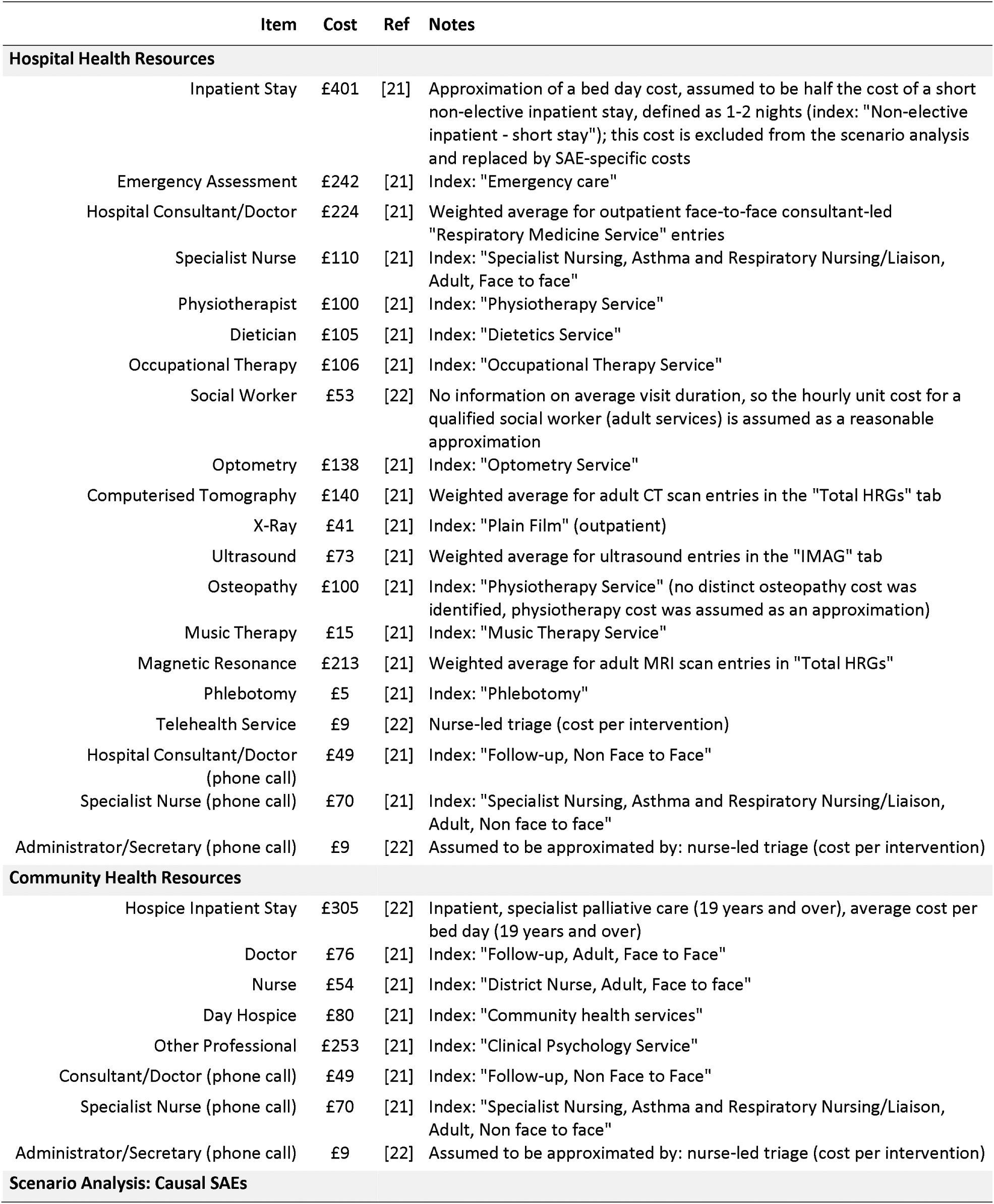
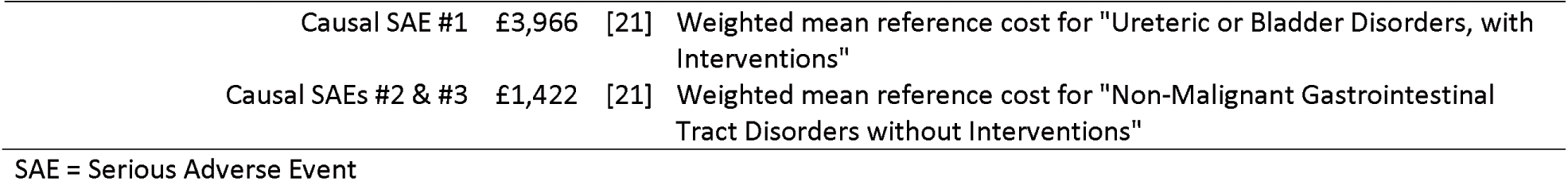
MABEL HRU Unit Costs.

In light of an observed imbalance in hospital admissions deemed by the investigator to be unrelated to morphine, a *post hoc* costing scenario was undertaken in which hospitalisation costs were measured only by Serious Adverse Events (SAEs) using industry-standard definitions considered to be related to morphine by site clinicians and study investigators, instead of the number of self-reported inpatient days which formed the base case *a priori*. All SAE reporting (including causality) was done whilst still blinded, except one SAE that was judged as having a causal relationship *post hoc*; this increased the number of causal SAEs (all in in the morphine arm) from 2 to 3, leading to a more conservative result than the fully blinded causality. In the scenario analysis, the hospital stay data were collected from the SAE reporting in the case report forms which were verified from the clinical record by site investigators and study monitors, as opposed to inpatient stay data which were sourced from patient questionnaires. Corresponding unit costs were sourced and approved for the three SAEs included in the scenario analysis by the Chief Investigators (reported in Table 1).

Per-patient costs were calculated by multiplying each patient’s recorded HRU units by their corresponding price weight.

All costs are reported in British Pounds (GBP) for the financial year of 2021/22, based on the most recent NHS reference costs available at the time of conducting the cost analysis. [21]

### Analysis

To account for missing data and the non-normal and skewed distribution of estimates, statistical tools from the validated *bootImpute* R programming package were used to estimate cost and QALY 95% confidence intervals (CIs) for means and differences in means between trial arms. [23,24] These methods combined multiple imputation by chained equations, non-parametric bootstrapping, and generalised linear model regression with a Gamma distribution and a log link to account for estimate skewness. [23,25,26]

The regression model, consistent with the main clinical trial analysis, controlled for age, sex, study site, and disease. [8] QALYs were additionally adjusted for baseline HSUV, as recommended by Manca *et al.* [17]

Estimates of the regression analyses from the simulated bootstrapped datasets and estimates from the pooled regression model were used to complete the cost-effectiveness analysis. As per NICE guidelines, the primary cost-effectiveness metric was the ICER measured in incremental costs per QALY gained (intervention minus control). The ICER was calculated alongside measures of parameter uncertainty in the form of a Cost-Effectiveness Plane (CEP) scatterplot and a Cost-Effectiveness Acceptability Curve (CEAC). [14]

### Concomitant medications

In contrast to the other patient-reported variables, concomitant medication data were collected from a CRF. Its analysis proved challenging for the following reasons:

- The concomitant medications CRF had a large number of entries (n = 1,852)
- Drug names were recorded as a free text variable, prone to erroneous and inconsistent spelling
- Many values were impossible to interpret (e.g., dose frequency recorded as “Other”)
- Each entry needed to be cross-referenced with a unit cost from the BNF
- many concomitant medications were unrelated to the MABEL trial (e.g., unrelated to breathlessness or morphine-related side effects)

To resolve these issues, the following steps were taken: the 1,852 concomitant medication data entries were cleaned, from which 424 unique categorical drug values were extracted (including a missing/error term); then, the 424 drug names were classified into drug categories by a clinician and health economics lead on the MABEL trial team, 328 of which were classified as ‘not relevant’ and were therefore excluded from cost calculation. Then, the unit costs of the remaining 96 drugs were identified by a web-scraping algorithm written in R with access to the BNF database. Then, the observed medication cost was calculated for each non-excluded entry (n = 444 out of N = 1,852) by multiplying the unit cost by the units used (as recorded in the CRF). At this point, there was still too much missing data to sum up medication costs by the patient, so the remaining data were imputed and bootstrapped using *bootImpute* in parallel to the patient-level data. This allowed imputed total per-patient concomitant medication costs to be calculated separately for each iteration of the simulated dataset.

### Patient and public engagement

No additional patient and public involvement was undertaken specifically for the trial’s health economic evaluation, beyond that of the main trial (see main clinical trial report for details). [8]

## RESULTS

### Study population

140 participants formed the MABEL intention-to-treat population (see Table 2). The CONSORT (Consolidated Standards of Reporting Trials) flow diagram (reporting checklist) is provided in Supplementary Figure 1. The main clinical paper describes further details of the screening and randomisation processes. [8]

**Table 2:**
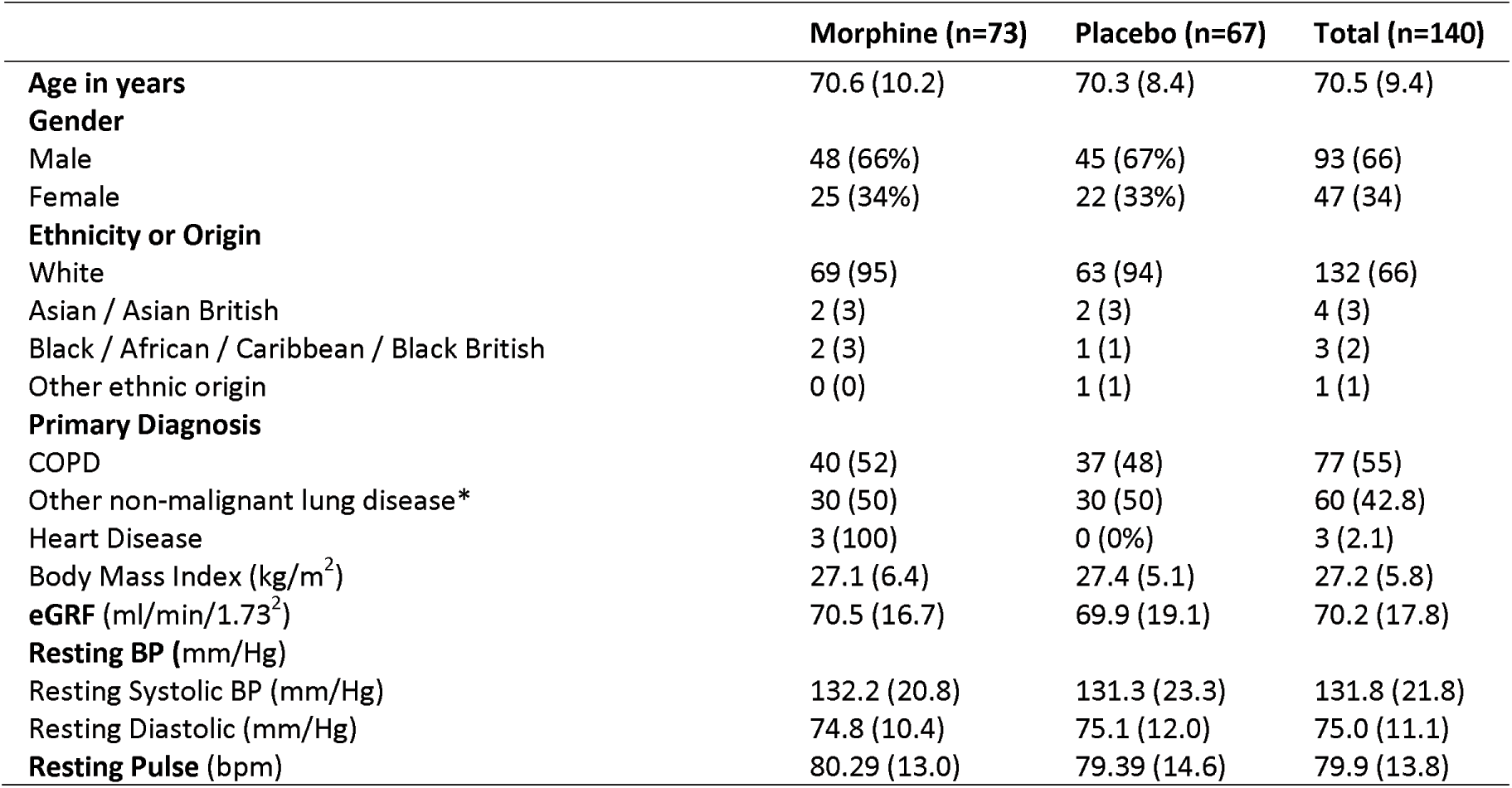
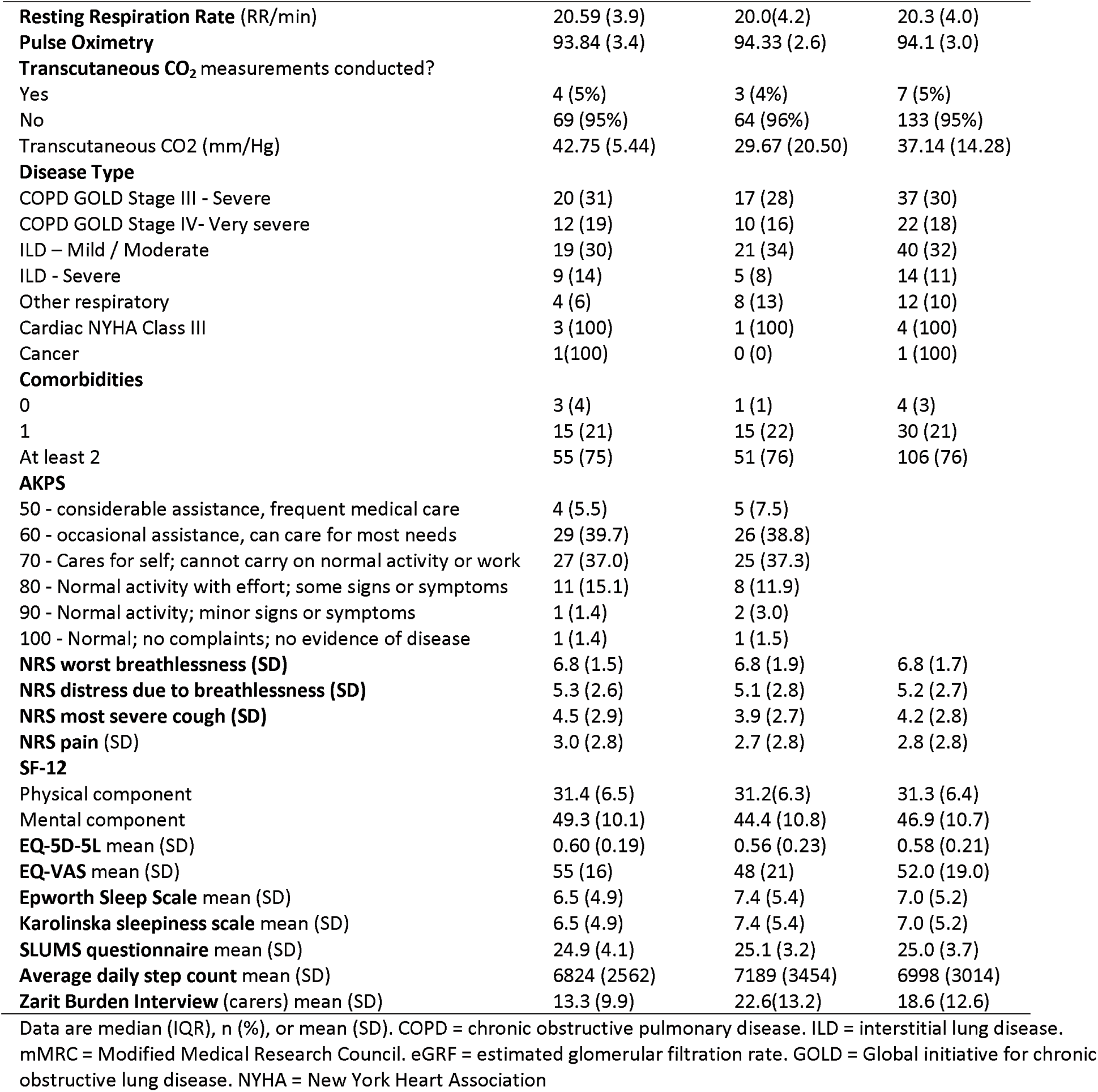
Baseline Characteristics.

Deaths over time by study group are presented in Figure 1 and, in conjunction with EQ-5D-5L data, were used to calculate QALYs.

**Figure 1:**
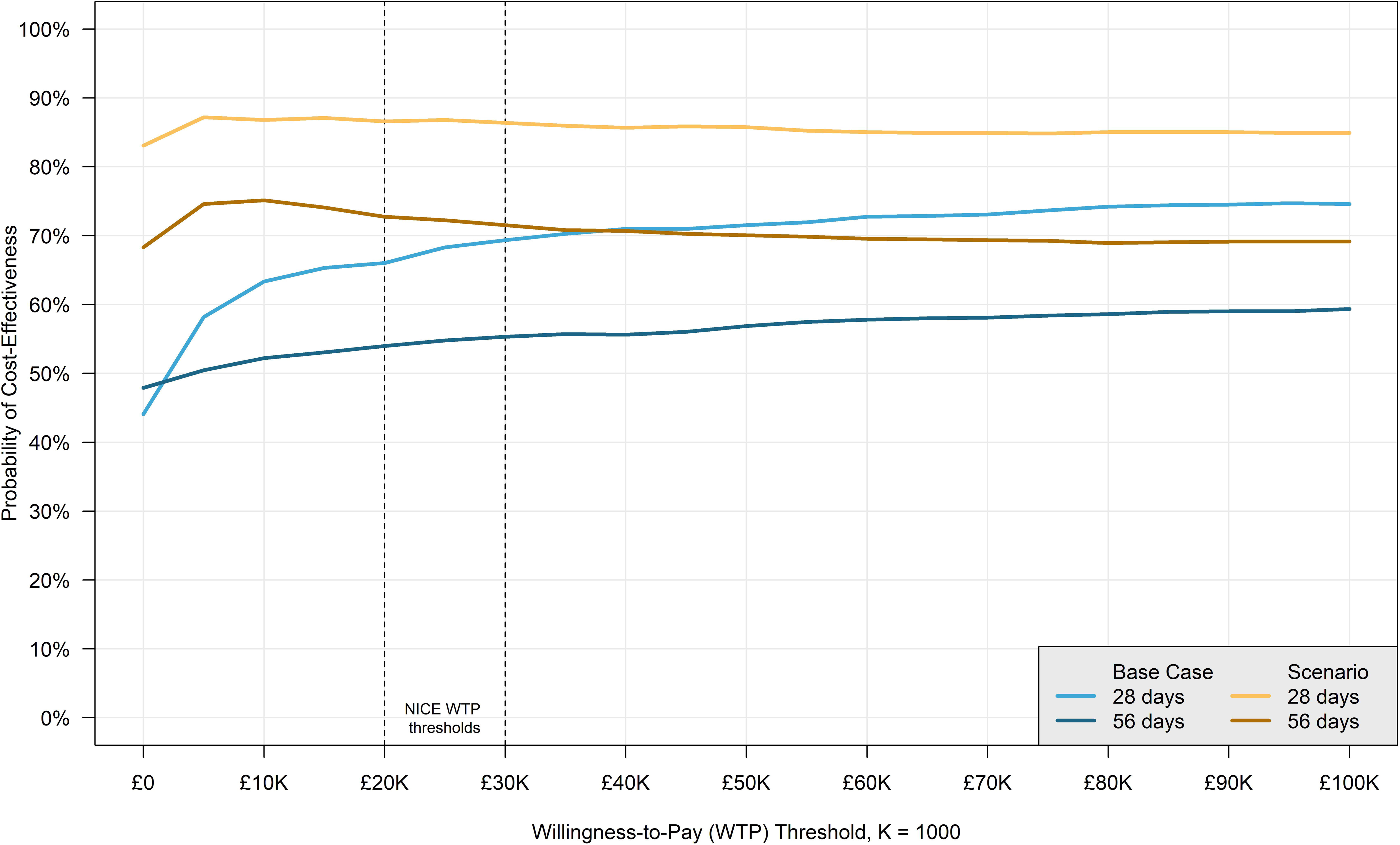
Mortality over Time by Study Group

### Data quality

The percentage of patients in each arm with missing values is categorised by each patient-reported health economic variable and presented in Supplementary Table 2. In these variables, missingness was below 20%. Unsurprisingly, missingness was highest for variables collected on Day 56 since randomisation, followed by those gathered on Day 28, and 0% for baseline measures. For both health outcomes and resources/costs, missingness is consistently slightly higher in the morphine arm (by up to 9% points).

Concomitant medication data is summarised separately in Supplementary Table 3. The table shows the number of entries in the CRF and the number and percentage of receiving patients by concomitant medication category.

### Health resource use

Table 3 shows the sums of patient-reported health resources used by each study group, which were used to calculate hospital and community costs.

**Table 3:**
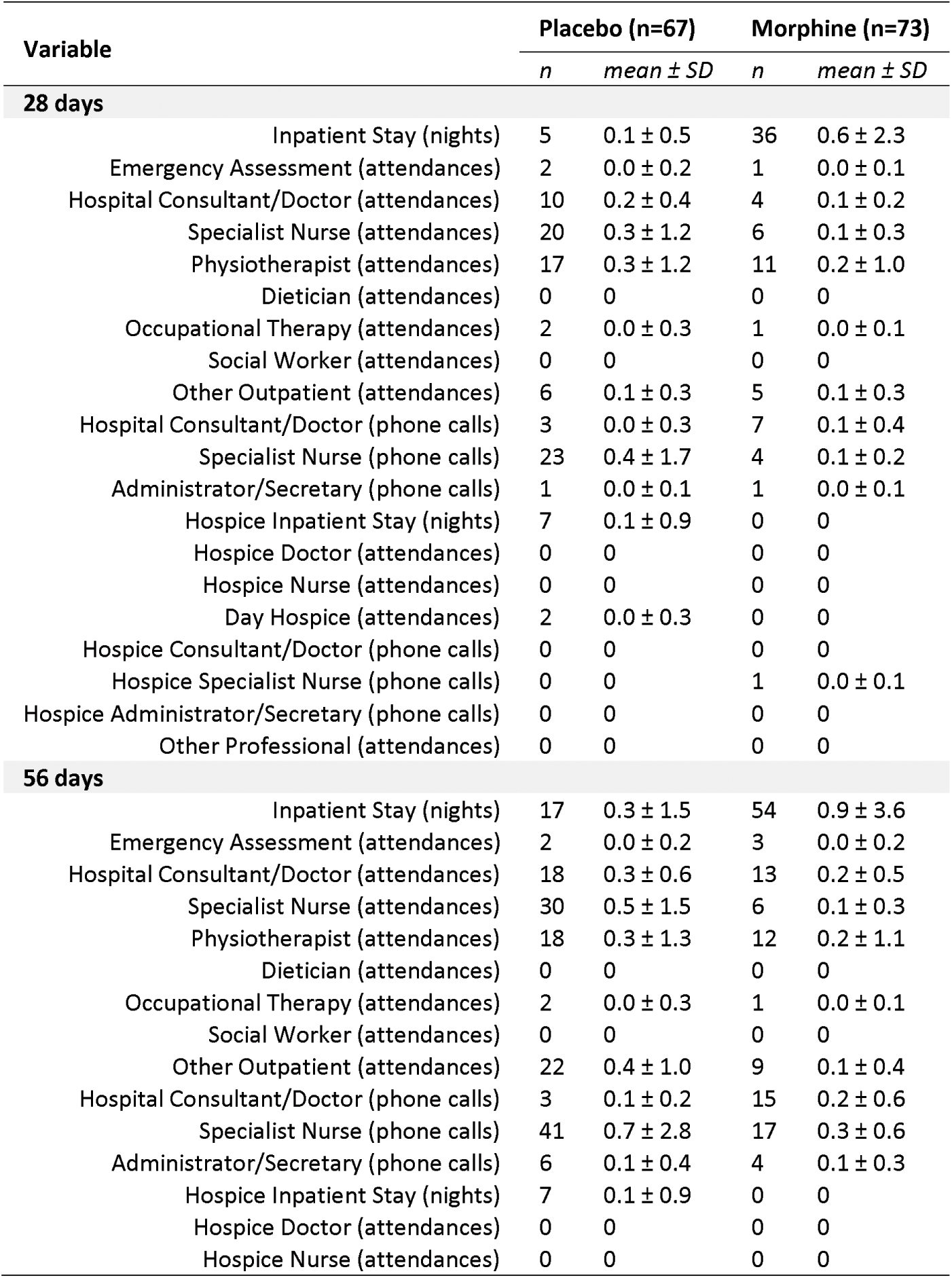
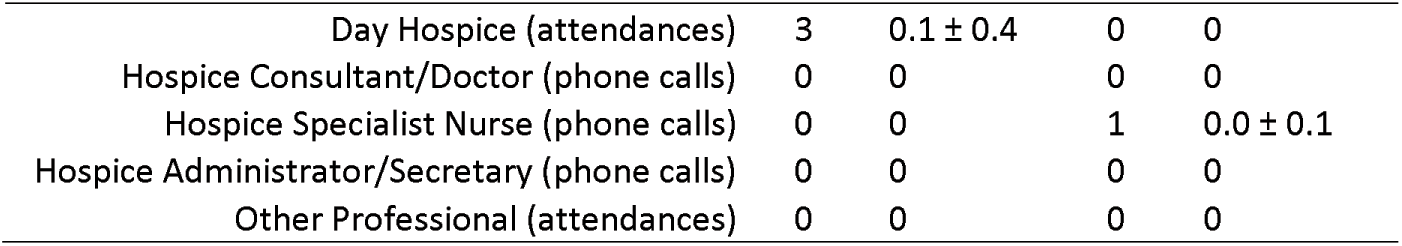
Health Resource Use by Study Group - n (summed total by study group), mean ± SD.

Table 3 shows a disparity in resource use patterns; patients in the Morphine arm spent more inpatient nights than Placebo (54 vs 17 by day 56), while the Placebo arm used more outpatient and community services. While this could be explained by random variation, it is also possible that due to small numbers, an imbalance in unrelated non-intervention-related SAEs has led to a spuriously high admission rate in the intervention arm. Therefore, we conducted a *post hoc* scenario analysis, wherein the self-reported inpatient stay nights were replaced with SAE hospitalisations recorded in the CRFs that were likely related to the intervention; three such SAEs were identified amongst three patients in the Morphine arm (one due to urinary retention, and two due to constipation) and 0/3 SAEs in Placebo.

### Regression analysis results

The summary statistics of the main MABEL economic variables are reported in Table 4. The *Placebo* and *Morphine* arms present mean per-patient values and 95% CIs estimated via non-parametric bootstrapping. The *Difference* column presents regression estimates and 95% CIs of the mean effects of the IMP on each variable, adjusted by the regression controls.

**Table 4:**
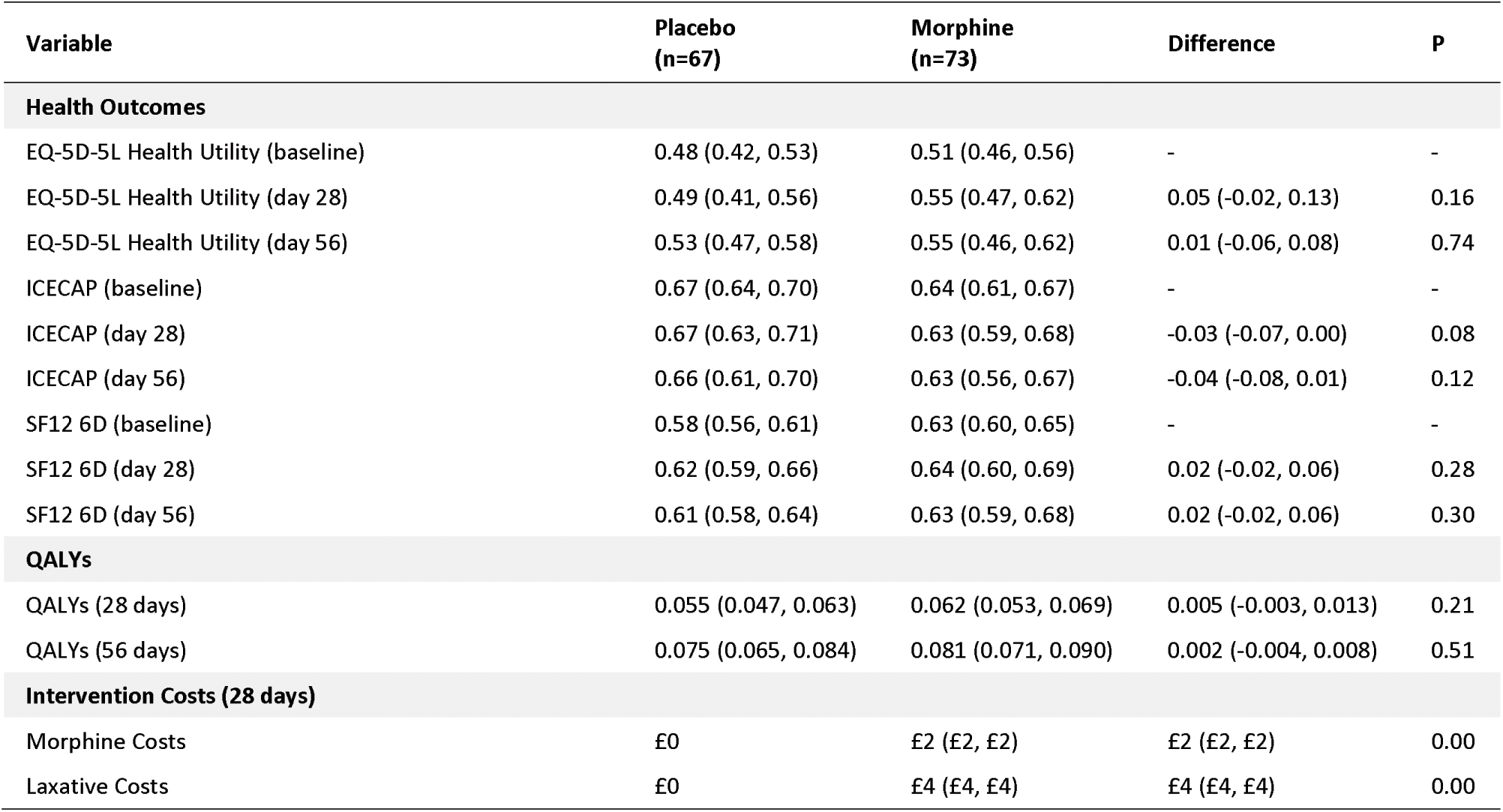
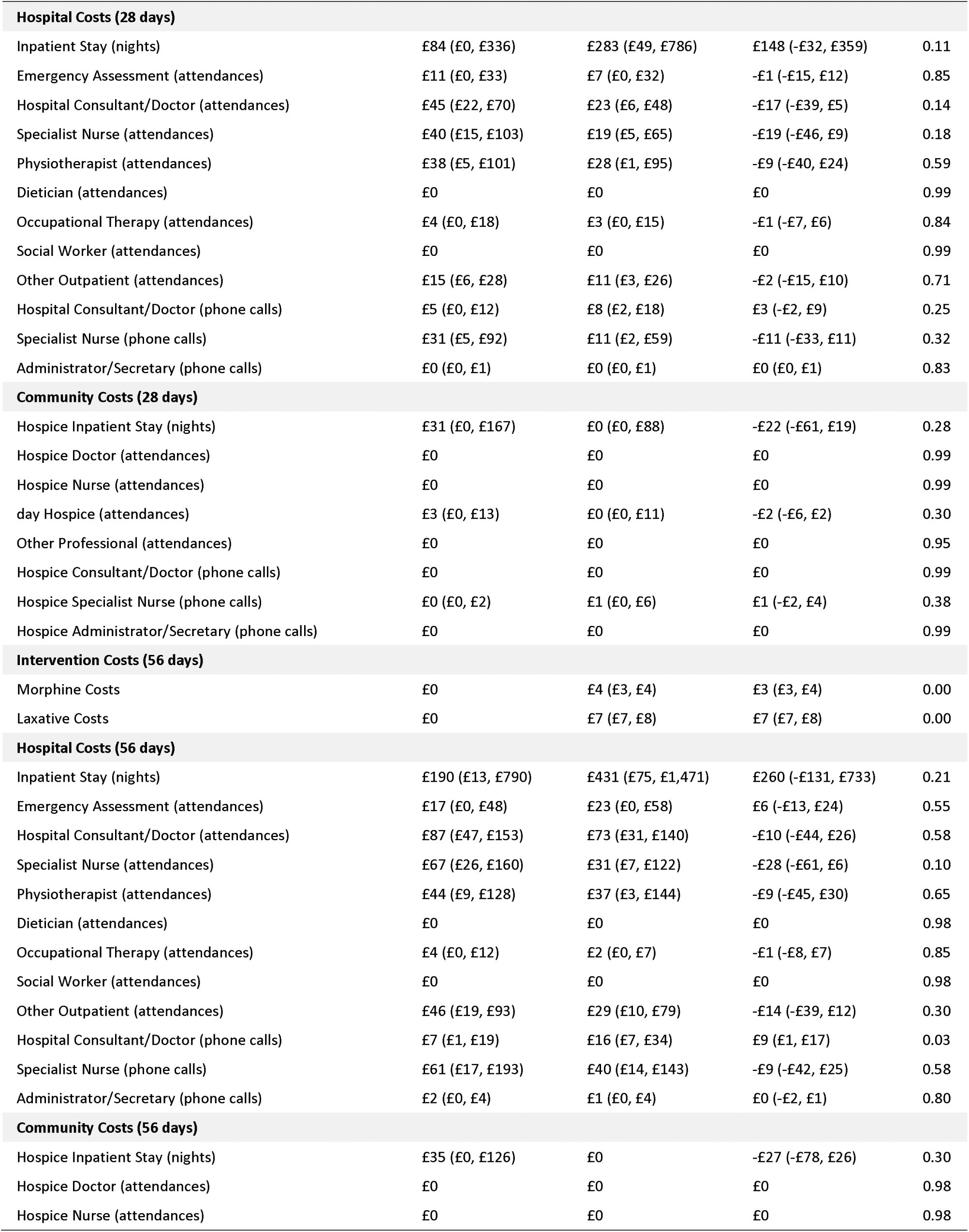
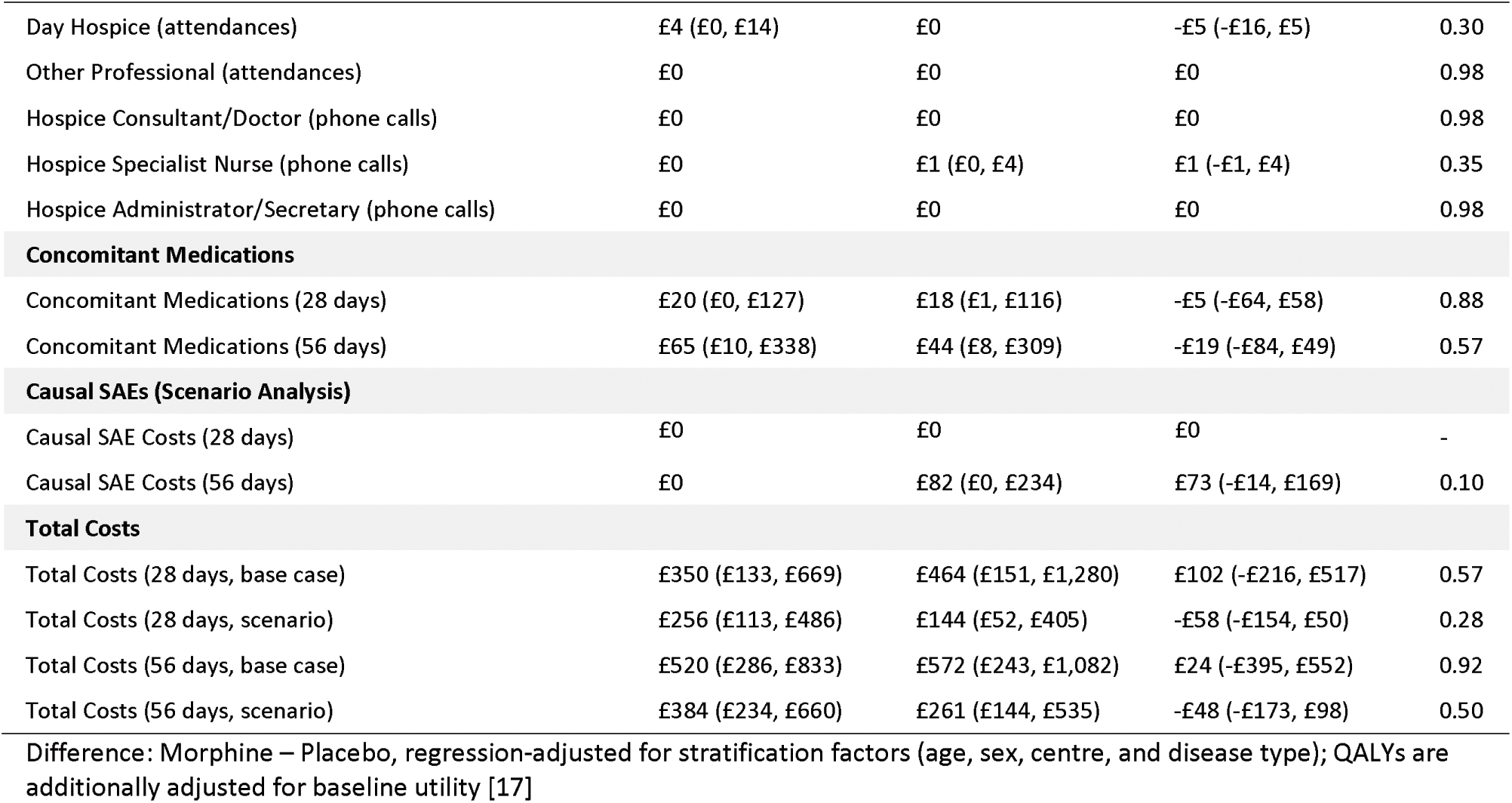
Summary Statistics, mean estimate (95% CI)

Overall, Table 4 presents no statistically significant differences in health outcome measures or costs between trial arms, with only two exceptions:

1. IMP and NIMP costs, which are set to £0 in the Placebo arm but are £4 (95% CI: £3, £4) and £7 (95% CI: £7, £8) in the morphine arm, respectively;
2. Hospital Consultant/Doctor phone calls have a small but statistically significant (P = 0.03) difference, being £9 (95% CI: £1, £17) higher in the Morphine arm relative to Placebo.

### Cost-effectiveness analysis results

The 56-day ICER for the MABEL intervention was estimated as a function of the adjusted difference in QALYs and total costs, resulting in 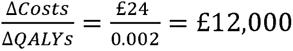 per QALY gained in the base case. In the sensitivity analysis, Morphine lowers costs by -£48 (95% CI: -£173, £98), therefore the ICER cannot be calculated and the intervention dominates standard care.

Uncertainty around the ICER results is presented in the Cost-Effectiveness Plane (CEP) in Figure 2, which shows the percentage of simulated datasets found within an area of the graph. For comparison, the base case and sensitivity analysis results were plotted together, therefore the standard CEP scatterplot was not used to improve readability and instead replaced with data ellipses plotted using the *dataEllipse* function from the *car* library in R. [27] The distribution of simulated results is presented in Table 5 as a percentage of simulated results in each quadrant of the CEP.

**Figure 2:**
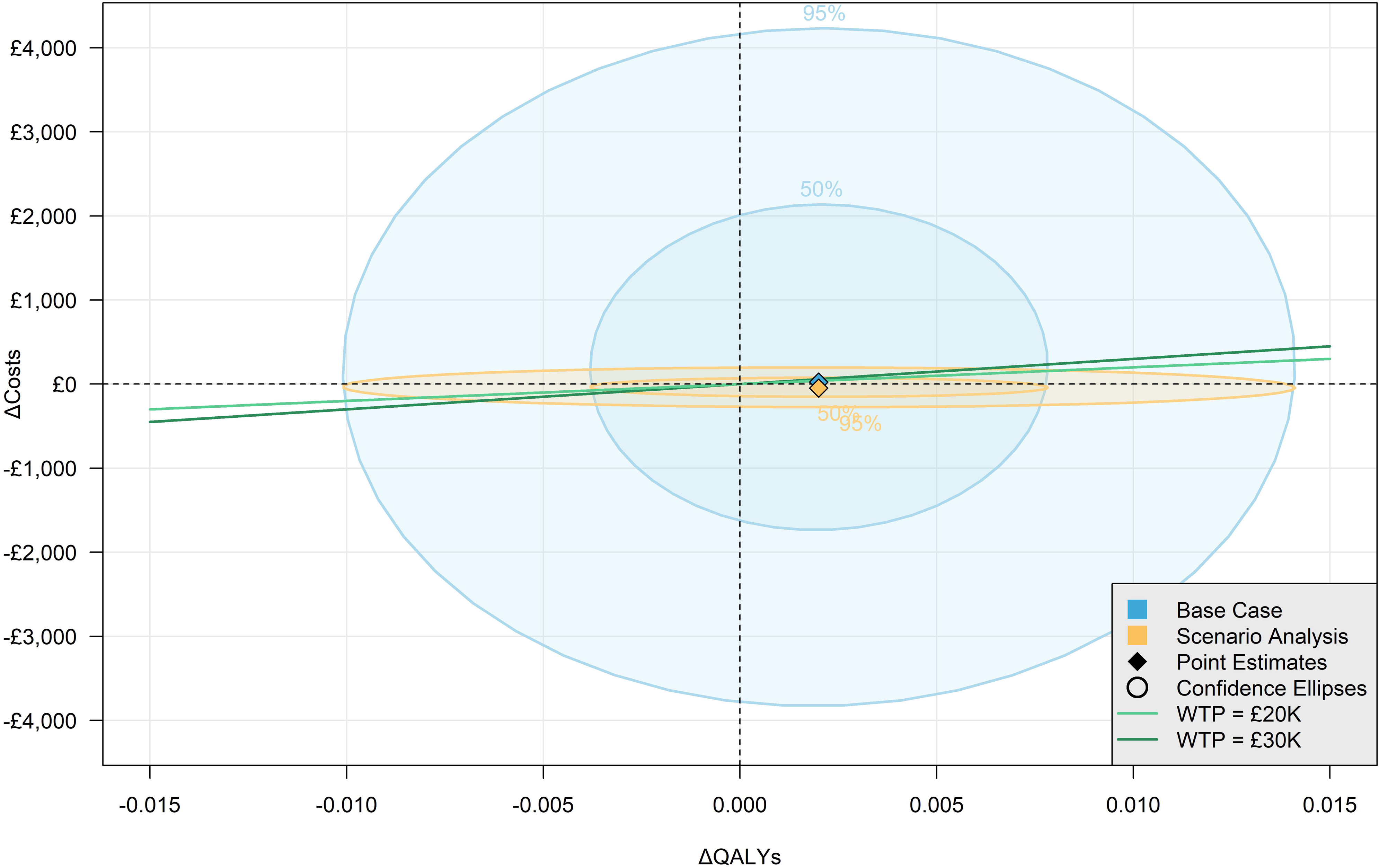
Cost-Effectiveness Plane

**Table 5:**
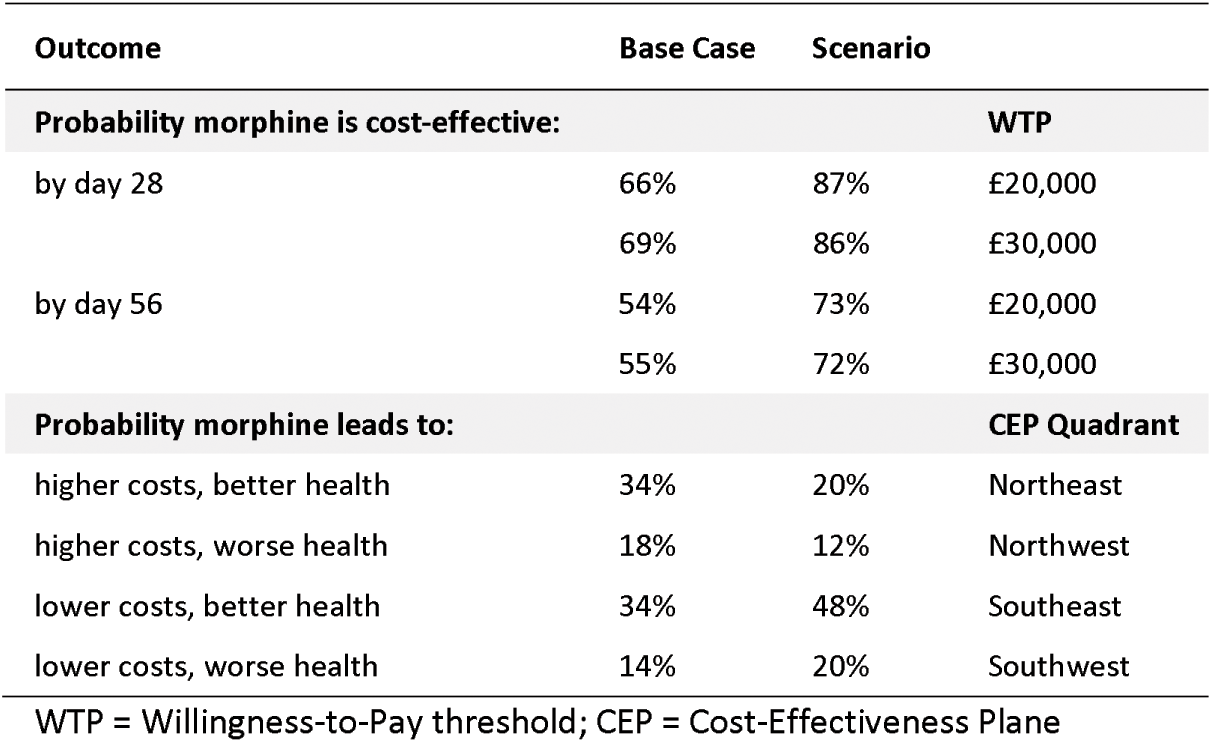
Summary of Cost-Effectiveness Results.

The Cost-Effectiveness Acceptability Curve (CEAC), which plots the probability of morphine’s cost-effectiveness compared to standard care against a WTP threshold, is presented in Figure 3. The probabilities of cost-effectiveness using NICE WTP thresholds of £20,000 and £30,000 are present in Table 5. A Value of Information (VoI) analysis was conducted alongside the CEAC calculations. The VoI results are presented in Supplementary Figure 2, which shows the Expected Value of Perfect Information (EVPI) plotted against the WTP threshold.

**Figure 3:**
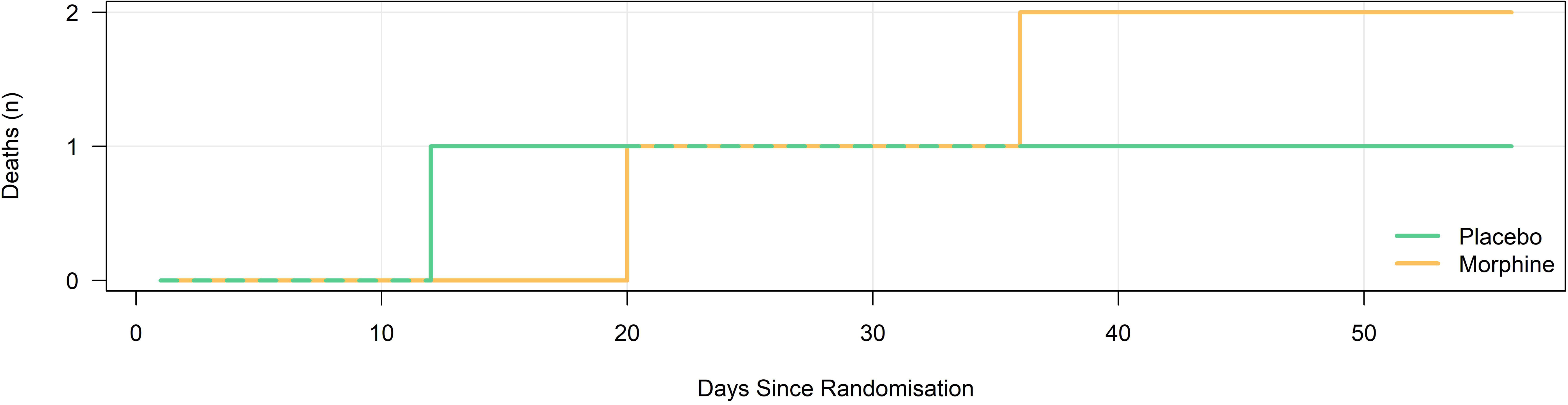
Probability of Cost-Effectiveness

## DISCUSSION

The analysis suggests that long-acting morphine may be cost-effective for chronic breathlessness, at a probability consistent with the previous health economic evaluation in the context of COPD. [7] In the scenario-based *post hoc* analysis, the probability of cost-effectiveness becomes greater. However, the adjusted results of MABEL’s health economic analysis show no overall statistically significant difference in costs or health outcomes between the placebo and long-acting arms and the cost-effectiveness estimates are highly uncertain. This is consistent with its main clinical results, which show that long-acting low-dose, oral morphine had no statistically significant effect on patient-reported worst breathlessness in people with chronic breathlessness. [8] The ICER is difficult to interpret when taken in isolation due to the small sample size and effect of a likely random imbalance in deaths, therefore we suggest that interpretation should focus on cost consequences analysis as this allows us to unpick possible relationships between adverse events, morphine, costs and patterns of health service use.

Despite the lack of statistical significance in the cost analysis, an interesting divergence in patient-reported HRU patterns was observed between trial arms. More outpatient services were used in the placebo arm, while the Morphine arm patients used more inpatient services. These inpatient admissions appeared to be imbalanced and unexplained by adverse events plausibly related to morphine. The scenario analysis proved a useful tool to rebalance this in case the trial’s sample size exacerbated the imbalance. However, a limitation of the scenario analysis stems from it being conducted *post hoc* and judging one of the SAEs as causal only after unblinding, unlike the fully double-blinded base case analysis. However, observational studies of cohorts of people with advanced COPD and interstitial lung diseases (conditions that 55% and ∼40% of the MABEL trial population had respectively) demonstrated no excess hospital admissions or deaths in people taking opioids at less than 30mg morphine-equivalent doses per day. [28,29] This is despite having very severe respiratory disease; all participants in these cohorts being on long-term oxygen therapy. Of note in the MABEL trial, where hospital admission was attributed to morphine, two of the three admissions were related to constipation. This emphasises the importance of close attention to bowel care in anyone taking morphine – this is a potentially avoidable situation, and in retrospect, the blinded laxative in the MABEL trial should have included a stimulant as well as a softener. Further, we need to consider whether some infective adverse related events might be related to morphine. There is a theoretical concern about the impact of opioids on the immune system, although the effect on clinical outcomes is unknown. However, we found no evidence that participants in the morphine arm experienced more infective events than the placebo arm, making it unlikely that relatedness was misclassified with this regard. [8]

The MABEL analysis of health outcomes produced some unexpected results. Adjusted point estimates for morphine’s impact on EQ-5D and SF12 scores on day 56 are positive, while those for ICECAP are negative. While not statistically significant, this is still the opposite trend to what we expected before data analysis, as it suggests that participants taking morphine are *less* capable. However, this result could be consistent with the HRU analysis, wherein morphine arm participants used more inpatient services (e.g., hospitalisation) and fewer outpatient services relative to placebo. Hospital in-patient stays would be expected to impair capability, although this is not connected to and does not suggest morphine-related cognitive bluntness. The conflict with EQ-5D and SF12 scores might stem from the pain reduction morphine provides, but the clinically and statistically significant improvement in pain in the morphine arm at Day 7 (mean difference -0.92; 95% CI: -1.70, -0.13; p = 0.2) was not sustained throughout the 56 days of the MABEL trial other than a small and insignificant improvement. [8] Undoubtedly the contrasts between these health outcomes highlight the importance of future research to find the most appropriate measures of health outcomes in the context of advanced illness. It is also interesting to note the cost implications of out-patient and community care. People with moderate to severe breathlessness due to chronic medical conditions are high users of all aspects of healthcare (community, emergency, hospital) with costs also driven by community care health service needs. [6]

We highlight several strengths and weaknesses. While the robustness of patient-reported data collection was a significant strength of the analysis, the difficulties in analysing data from concomitant medications CRF proved to be a non-trivial study limitation. Future trials based in the UK could avoid these issues by ensuring drug names are not entered as free text and instead selected or searched from a pre-specified list corresponding to entries in the BNF; this would minimise errors created by spelling or data entry mistakes and ensure observed drugs can be matched with the price weights needed to estimate medication costs. Furthermore, CRF design could ensure a more consistent way of recording the dosage to let researchers easily identify how many units of a given drug each patient has taken over the study time horizon. The challenge in this patient population (nearly all had at least one co-morbidity), is that most are taking multiple medications.

There are significant limitations in the ICER analysis as described above. There are also challenges when health economics are applied within a palliative context. It is likely that if morphine was a new drug developed for the management of breathlessness, a licence would not be granted based on efficacy, making cost-effectiveness irrelevant. However, in the post-marketing context of morphine being *re-purposed* for use in chronic breathlessness, we estimate that if morphine was implemented for this indication, then, on balance, the use of morphine would improve net health gain from our current NHS budget. However, this does not necessarily mean it is the correct thing to do from a clinical point of view

In conclusion, while the MABEL health economic analysis does not show statistically significant effects of morphine on costs or health outcomes in people with chronic breathlessness, the scenario analysis shows a high probability of cost-effectiveness stemming from small non-statistically significant improvements in health outcomes and cost savings due to lower outpatient and community health resource usage. However, the ICER is difficult to interpret due to the small sample size and effect of a likely random imbalance in deaths, and interpretation should focus on cost consequences analysis. Despite these limitations, this study presents high-quality clinical trial results, contributes to the health-economic literature on morphine and breathlessness, and provides robust estimates that can be used in future economic modelling.

## Trial details

Trial registration ID: ISRCTN87329095

EudraCT number: 2019-002479-33

Recruitment dates: 01/03/2021 (start) - 31/10/2023 (end)

## Author contributions

Atter M (MA): conceptualisation, methodology, validation, formal analysis, data curation, data verification, data interpretation, writing (original draft), writing (review, critical input and editing), visualisation, software

Hall PS (PSH): conceptualisation, methodology, validation, funding acquisition, data curation, data verification, data interpretation, writing (review, critical input and editing), supervision; PSH acted as guarantor

Evans RA (RAE): contributions to clinical paper needed for the health economic analysis (data interpretation, writing, visualisation)

Norrie J (JN): funding acquisition, supervision

Cohen J (JC): funding acquisition, supervision, project administration

Williams B (BW): data curation, supervision, project administration

Chaudhuri N (CN): funding acquisition, data interpretation, writing (review, critical input, and editing)

Bajwah S (SB): funding acquisition, data interpretation

Higginson IJ (IJH): funding acquisition, data interpretation

Pearson M (PM): funding acquisition, data interpretation

Currow DC (DCC): funding acquisition, data interpretation, writing (review, critical input, and editing)

Stewart G (GS): patient recruitment

Fallon MT (MTF): conceptualisation, methodology, validation, funding acquisition, data interpretation, writing (review, critical input and editing), supervision, project administration

Johnson MJ (MJJ): conceptualisation, methodology, validation, funding acquisition, data curation, data verification, data interpretation, writing (review, critical input, and editing), supervision, project administration

## Data sharing

Data supporting this work are available upon reasonable request. Relevant stakeholders will review all requests based on the principles of a controlled access approach. Requests to access anonymised data should be made to.

## Acknowledgements

We thank the NIHR Research Delivery Network for their support of the study. Our thanks go to many individuals and organisations who contributed to this research (full list is included in the clinical paper’s supplementary material). [8]

## SUPPLEMENTARY MATERIALS

**Supplementary Table 1:**
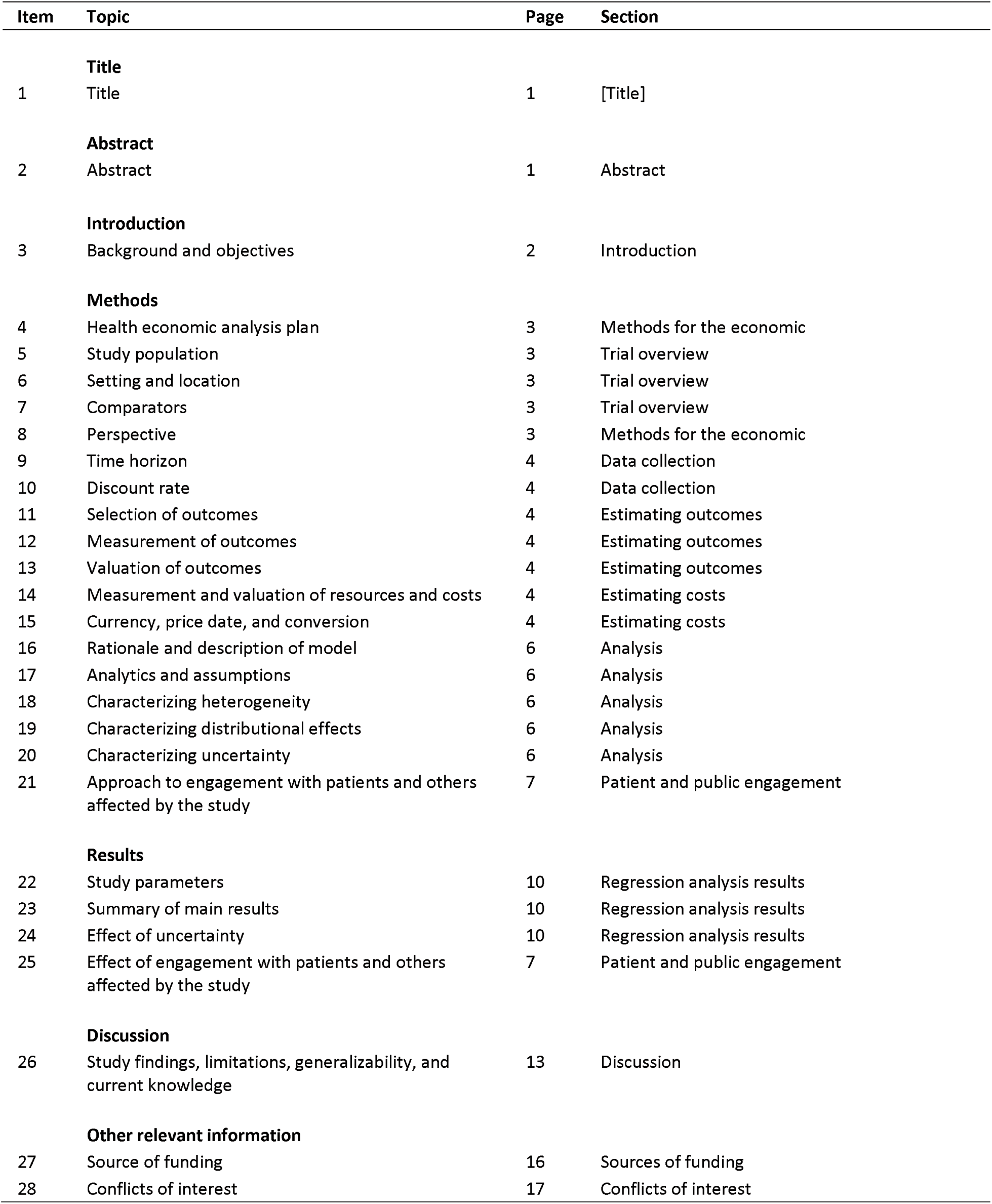
CHEERS (2022) Checklist.

**Supplementary Table 2:**
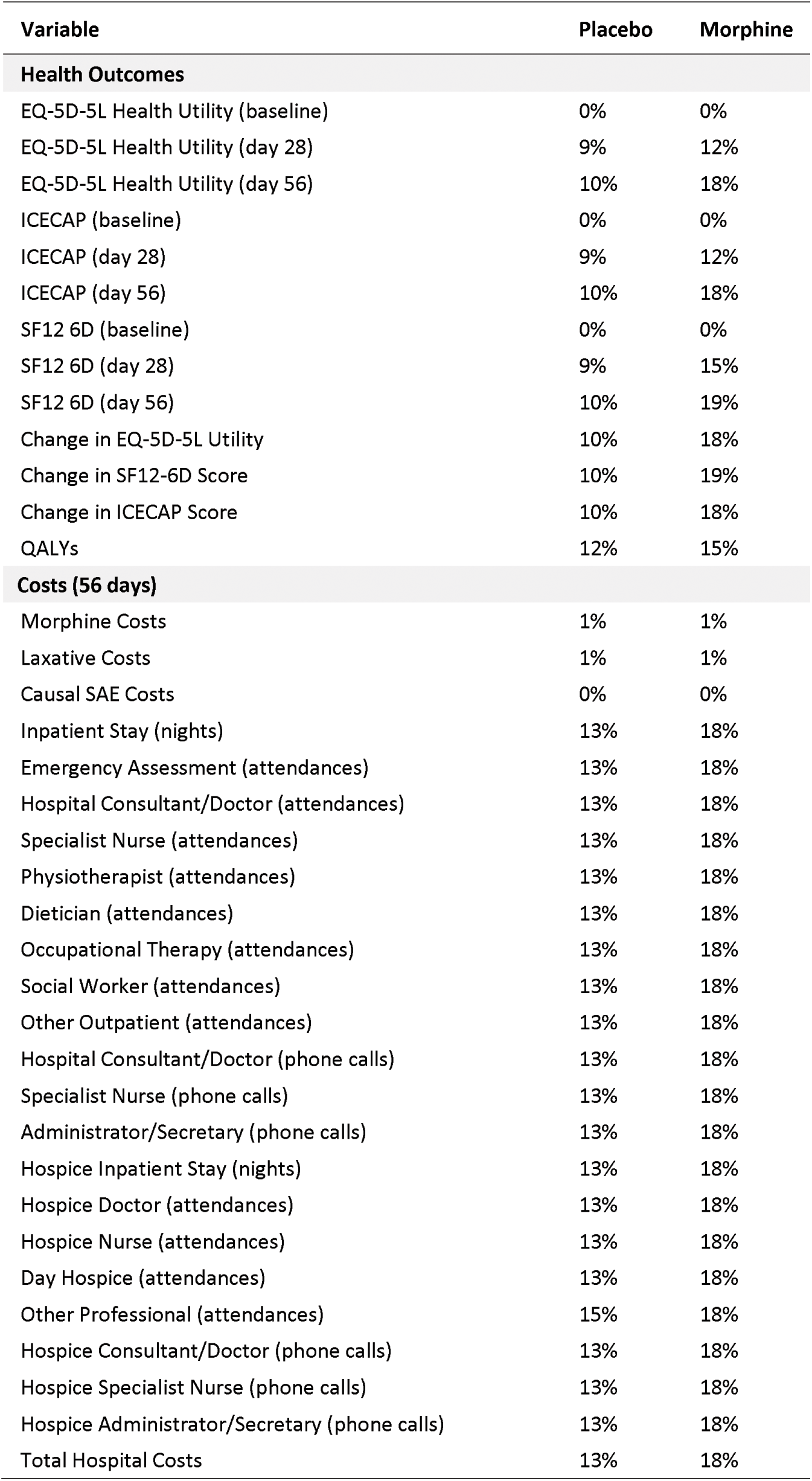
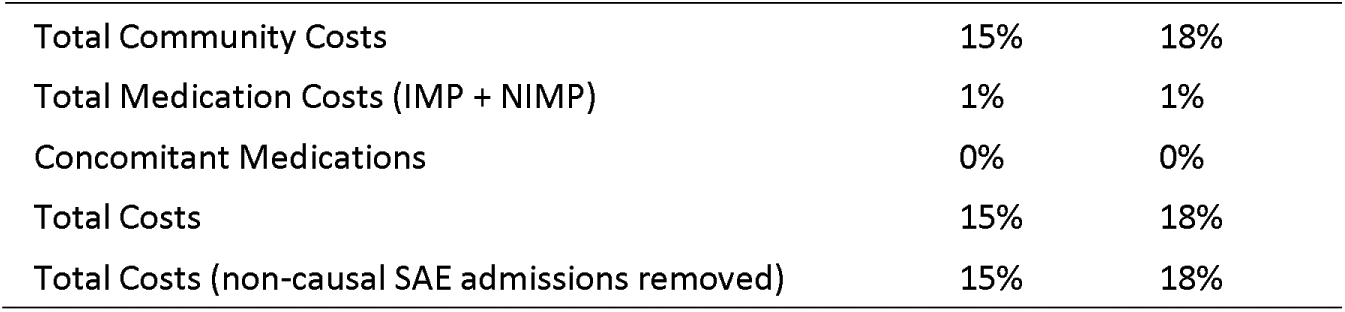
Percentage of Patients with Missing Data (Excl. Concomitant Medications)

**Supplementary Table 3:**
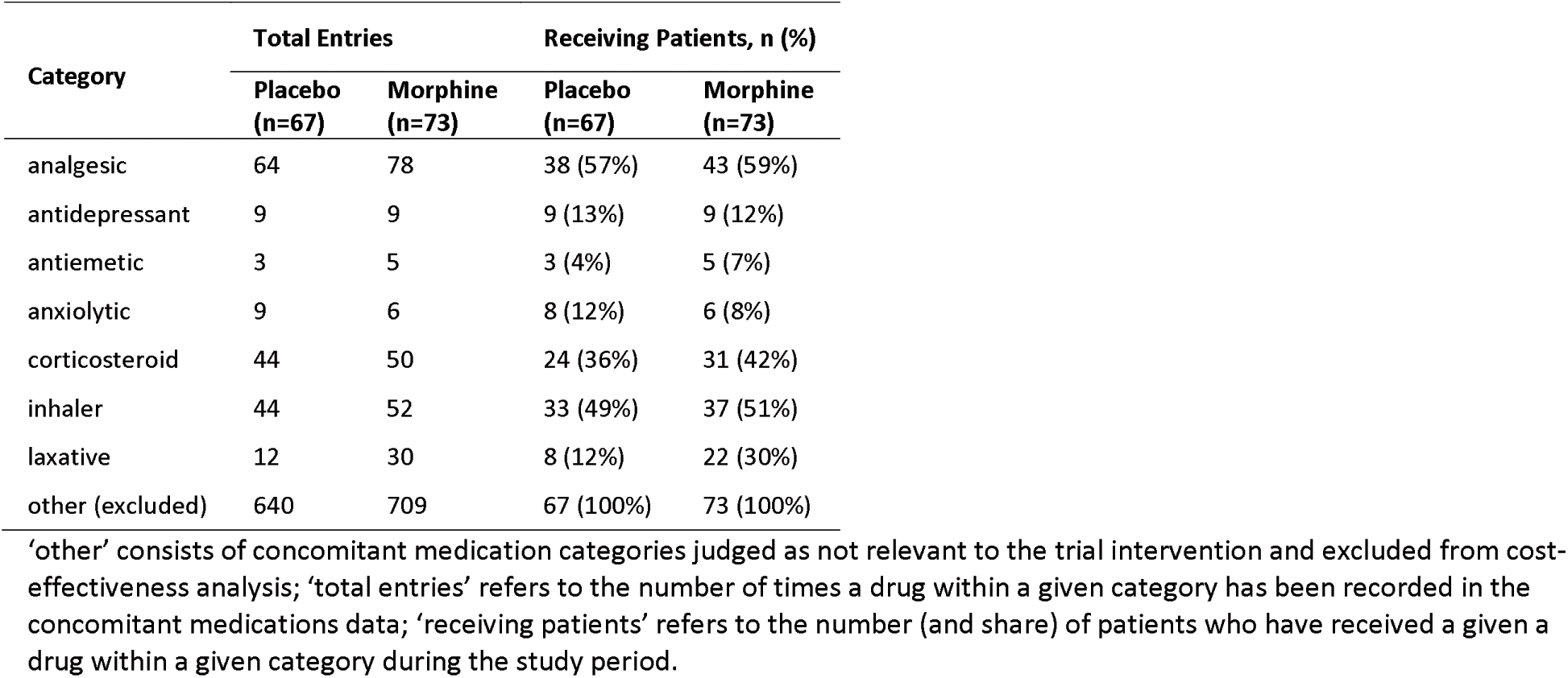
Unique Concomitant Medication Entries by Category.

**Supplementary Figure 1:**
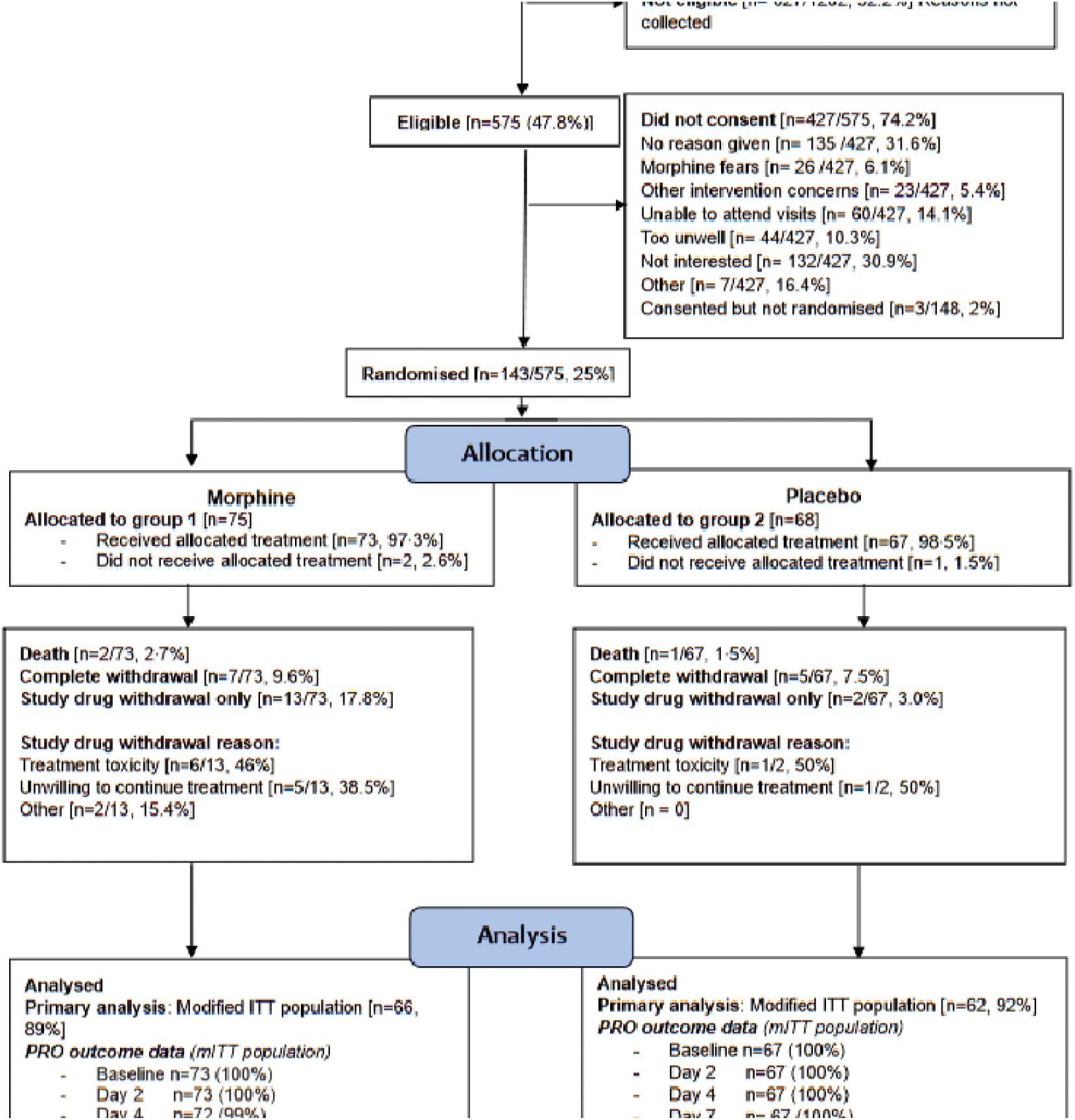
MABEL CONSORT Diagram

**Supplementary Figure 2:**
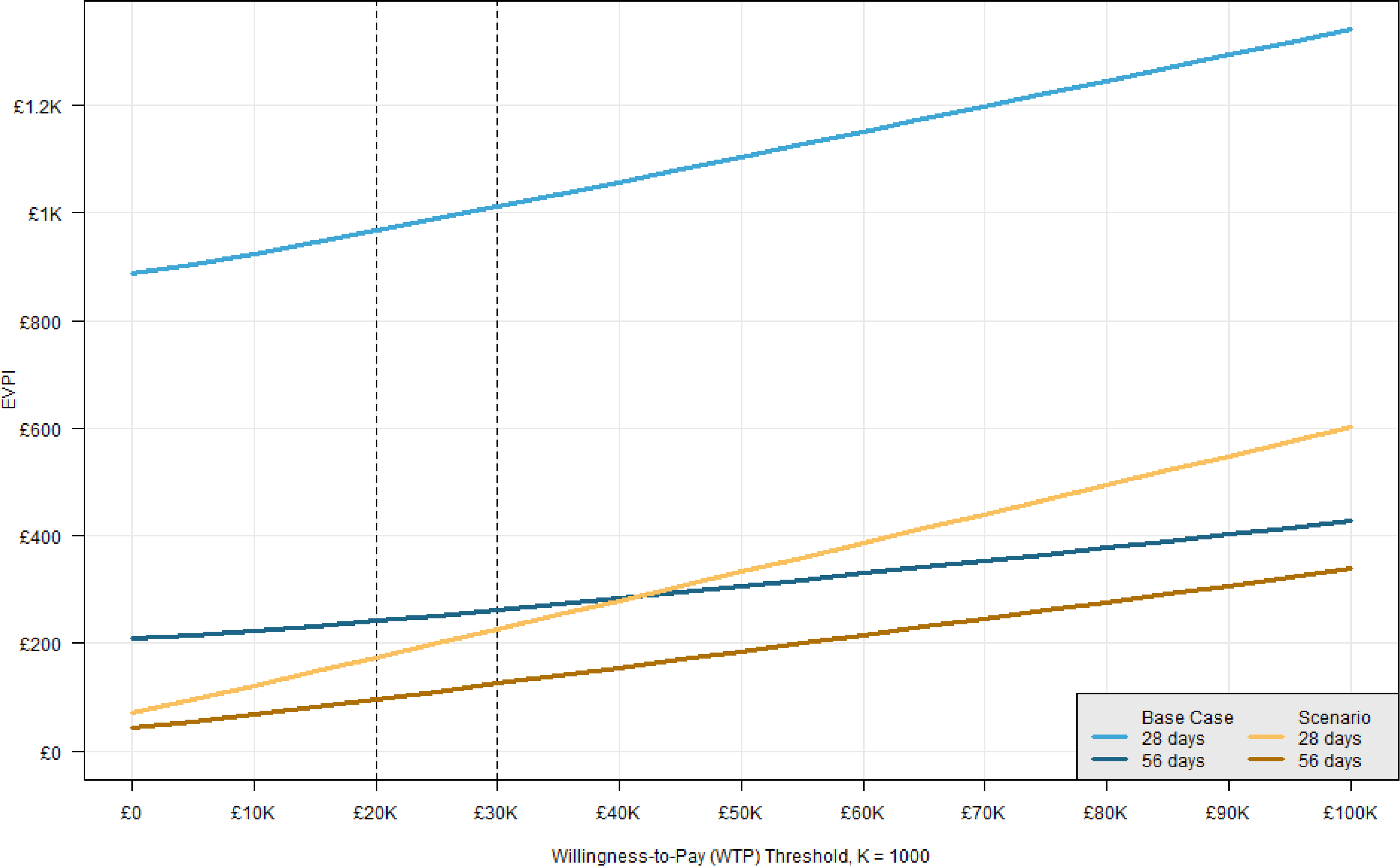
Value of Information Analysis: Expected Value of Perfect Information

